# Association of chronic kidney disease, ethnicity and socioeconomic status with COVID-19 hospitalisation and mortality: a UK Biobank study

**DOI:** 10.1101/2021.08.02.21261469

**Authors:** Thomas J Wilkinson, Courtney J Lightfoot, Alice C. Smith, Thomas Yates, Kamlesh Khunti, Francesco Zaccardi

**Affiliations:** Leicester Kidney Lifestyle Team, Department of Health Sciences, University of Leicester, Leicester, UK; Leicester NIHR Biomedical Research Centre, Leicester, UK; Leicester Diabetes Research Centre, Leicester, UK; NIHR Applied Research Collaboration East Midlands, Leicester, UK; Leicester Real World Evidence Unit, University of Leicester, UK

**Keywords:** COVID-19, ethnicity, health inequalities, socioeconomic status, chronic kidney disease

## Abstract

- In individuals with chronic kidney disease (CKD), Black and South Asian ethnic groups are twice as likely to have severe COVID-19 compared to White participants, whilst the most socially deprived groups are at a 50-60% increased risk of severe COVID-19.
- This study is the first to highlight the association between ethnicity and socioeconomic deprivation and the risk of severe COVID-19 among those with CKD in the UK.
- Interventions to reduce morbidity and mortality amongst these groups and policy and practice improvements are needed to address the broad disparity among CKD patients.

## Introduction

In the UK, as of July 2021, there have been over 4.5 million confirmed cases of coronavirus disease-2019 (COVID-19) and over 150,000 deaths caused by severe acute respiratory syndrome coronavirus 2 (SARS-CoV-2) [1]. Data are being reported on subpopulations most at risk of COVID-19 and its severest forms. Age, ethnicity, and socioeconomic position fundamental components in health inequality strongly influence health outcomes for both infectious and non-communicable diseases, and COVID-19 has further exposed the strong association between these and adverse health outcomes [2].

There is substantial evidence that a disproportionate impact of COVID-19 exists on Black and South Asian ethnic groups [3]. Individuals from these groups are more likely to be infected by SARS-CoV-2 and have an increased risk of intensive care admission compared to those of White ethnicity [4]. The mortality risk from COVID-19 among Black and Asian ethnic minority groups approximately twice that of White patients [2, 5-8]. Socioeconomic status is also a key factor in COVID-19 outcome [9, 10] and mortality rates from COVID-19 in the most deprived areas are more than double that of least deprived areas [6, 7, 11].

Many of the ethnic and socioeconomic disparities that increase susceptibility to COVID-19 also make individuals vulnerable to chronic kidney disease (CKD). The risk of CKD is higher in ethnic minority groups compared with White individuals at every CKD stage [12] and CKD is associated with greater hospitalisation and mortality from COVID-19 [7, 13-15]. Whilst the aetiology of CKD involvement is multifactorial [13], the interactions with ethnic and socioeconomic status have not been studied. Previous data suggests that inequalities in COVID-19 deaths by ethnic group exist among people with similar pre-existing conditions including CKD [7]; however, to our knowledge no study has investigated the role of ethnicity and socioeconomic status on COVID-19 severity among those with CKD, or whether existence of CKD increases risk in these groups further.

## Short Methods

### Data source

This study uses UK Biobank data. Participants from the general population were recruited between March 2006 and December 2010 [16]. UK Biobank was approved by the North West Multi-Centre Research Ethics Committee (11/NW/0382). All participants provided informed consent. This work was conducted under application number 52553.

### Population

Participants with CKD were identified by a creatinine-derived eGFR of <60 ml/min/1.73m^2^ and/or a urinary albumin/creatinine ratio (ACR) ≥3□mg/mmol (ACR A2-A3). Participants with an eGFR of ≥60 ml/min/1.73m^2^ and an ACR <3 mg/mmol were included as a non-CKD comparison group.

### Exposure

Our exposures of interest were ethnicity and socioeconomic deprivation status, obtained at baseline. Ethnicity was classified using Office of National Statistics groupings. To provide sufficient numbers for analysis, the analysis was limited to include White, South Asian, and Black ethnic groups; South Asian and Black individuals form the two largest minority ethnic groups within the UK. These groups have previously been used in UK Biobank analyses [8, 9, 17].

The Townsend deprivation index was used as a composite measure of socioeconomic status based on employment, car and home ownership, and household overcrowding; a lower value represents a higher socioeconomic status. Participants were classified into three groups: ‘least deprived’: less than -2.00; ‘average’: -2.00 to 1.99; and ‘most deprived’: ≧2.00 [18].

### Outcomes

Public Health England provided SARS-CoV-2 test results (available from 16^th^ March to 3^rd^ May 2021). Records were linked to inpatient Hospital Episode Statistics and national mortality registers (data available to 23^rd^ March 2021). As per previous studies [19], the primary outcome was ‘severe’ COVID-19 defined as a composite of a positive SARS-CoV-2 related hospital admission or death from COVID-19 (ICD-10 code U07.1/U07.2).

### Confounders

Covariates included: current age (calculated on date of SARS-CoV-2 test from original baseline assessment); sex; obesity (defined using body fat%); and number of cancer (including any incidence of cancer and leukaemia) and non-cancer reported illnesses (including cardiovascular disease and diabetes).

### Statistical analysis

Analysis was based on a whole population level approach [20], with severe COVID-19 cases compared to the remaining UK Biobank population. Maximally-adjusted complete-case logistic regression models were used to analyse the associations between ethnicity, socioeconomic status, and severe COVID-19. The results are reported as odds ratios (ORs) with 95% confidence intervals (95%CI). To investigate whether CKD modified the association between ethnicity/socioeconomic status and severe COVID-19 risk, a likelihood ratio test was conducted comparing two models, with and without an interaction term. Data was analysed using IBM SPSS (V26.0).

## Results

### Participant characteristics

Data were available for 459,042 participants, of whom 10,480 (2.3%) had CKD. In those with CKD, the median (25-75^th^ percentile) age was 76.0 (72.0-79.0) years, and 47.9% were males. The median eGFR was 54.2 (45.5-57.1) ml/min/1.73m^2^ . The majority of CKD participants were White (94.2%) (Table 1).

**Table 1.** Basic participant characteristics.

| N=459,042 | CKD | Non-CKD |
| --- | --- | --- |
| n (%) | 10,480 (2.3%) | 448,562 (97.7%) |
| Age (years) | 76.0 (72.0-79.0) | 69.0 (63.0-76.0) |
| Sex (male) | 5024 (47.9%) | 135,557 (43.4%) |
| *Ethnicity* |  |  |
| White | 9748 (94.2%) | 423,765 (95.7%) |
| Black | 345 (3.3%) | 8554 (1.9%) |
| South Asian | 198 (1.9%) | 1371 (0.3%) |
| *Townsend deprivation index* |  |  |
| Least deprived | 5194 (49.6%) | 234,945 (52.4%) |
| Average | 3279 (31.3%) | 141,498 (31.6%) |
| Most deprived | 1998 (19.1%) | 71,688 (16.0%) |
| eGFR (ml.min.1.73m<sup>2</sup>) | 54.2 (45.5-57.1) | 93.2 (82.8-100.0) |
| ACR (mg/mmol) | 2.0 (1.0-6.0) | 1.0 (0.0-2.0) |
| CKD Stage 3 | 8782 (95.2%) | - |
| CKD Stage 4-5 | 415 (4.5%) | - |
| No. of cancer illnesses | 0.0 (0.0-0.0) | 0.0 (0.00.0) |
| No. of non-cancer illnesses | 3.0 (2.0-5.0) | 1.0 (1.0-3.0) |
| Obesity | 6997 (69.5%) | 246,619 (56.2%) |
| *COVID-19 status* |  |  |
| Positive inpatient tests | 169 (1.6%) | 4338 (1.0%) |
| Death from COVID-19 | 78 (0.7%) | 953 (0.2%) |
| Severe COVID-19 | 247 (2.4%) | 5291 (1.2%) |

Data shown as median and inter-quartile range (25-75th percentile) for continuous and or n (%) for categorical variables. Cancer illnesses included bowel, skin, prostate, and leukaemia cancer; non-cancer illnesses included cardiovascular disease, respiratory conditions, diabetes, and neurodegenerative disease

There were 5538 cases of severe COVID-19: 247 (2.4%) occurred in participants with CKD and 5291 (1.2%) in those without (Table 1). Participants with CKD were 1.6 times more likely to have had severe COVID-19 (OR: 1.58; 95%CI: 1.38-1.82; P<0.001).

### Ethnicity and risk of severe COVID-19 in those with CKD

Differences in the crude frequency of severe COVID-19 cases across ethnic groups are depicted in Figure 1: overall, in participants with CKD, severe COVID-19 was more prevalent across all ethnic groups compared to White participants. In participants with CKD, compared to White group, Black ethnic individuals were approximately 2.3 times more likely to have had severe COVID-19 (OR: 2.26; 1.34-3.80; P=0.002) and those of South Asian ethnicity 2.1 times more likely (OR: 2.07; 1.07-4.00); P=0.032; Figure 2). A similar pattern of association was observed in participants without CKD, with no evidence of an interaction between presence of CKD and ethnicity (P=0.363; Figure 2).

**Figure 1.**
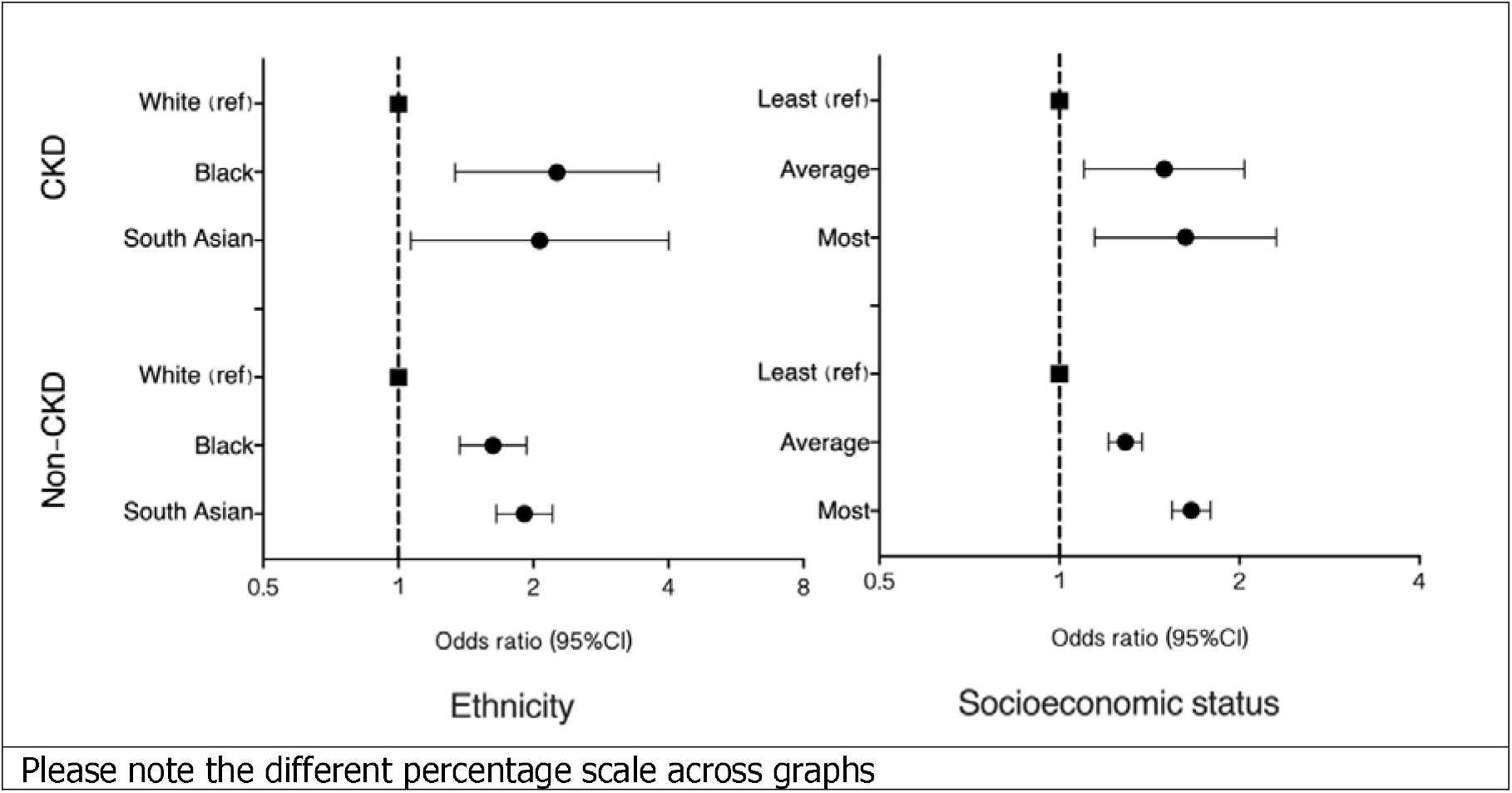
Proportion of COVID-19 positive inpatient tests, deaths, and severe COVID-19 across ethnic and socioeconomic deprivation groups, in participants with and without CKD.

**Figure 2.**
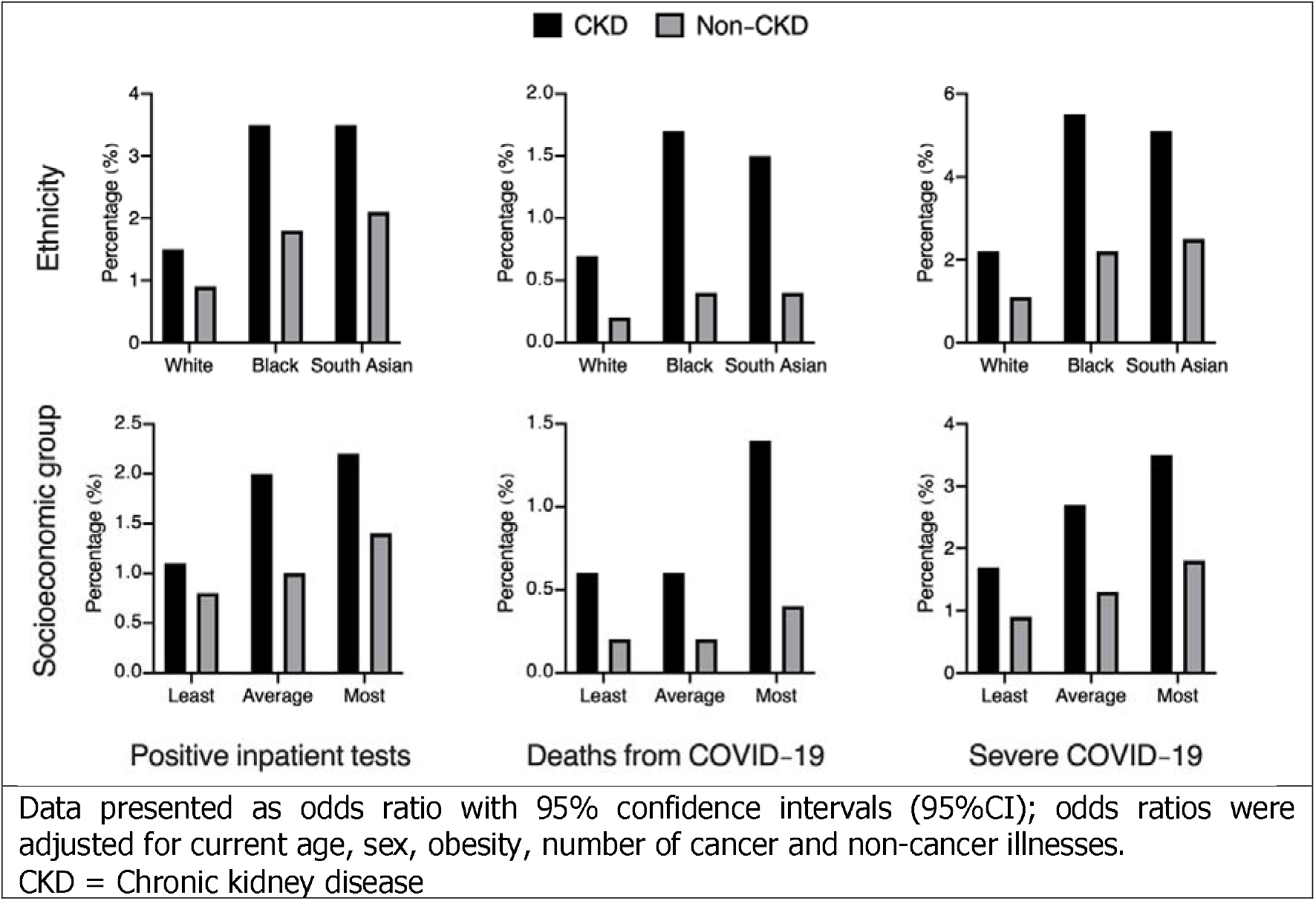
Likelihood of severe COVID-19 across ethnic and socioeconomic deprivation groups, in participants with and without CKD.

### Socioeconomic deprivation and risk of severe COVID-19 in those with CKD

Severe COVID-19 was more prevalent in participants with vs without CKD across all socioeconomic groups and was progressively more common in groups with a greater deprivation (**Figure 1**). In participants with CKD, compared to the least deprived group individuals in the most deprived group were 63% more likely to have severe COVID-19 (OR: 1.63; 1.15-2.31; P=0.007), whilst those in the average group were 50% more likely to have severe COVID-19 (OR: 1.50; 1.10-2.04; P=0.011; **Figure 2**). There was no evidence of an interaction between CKD and socioeconomic status (P=0.559).

## Discussion

In a cohort study of almost 500,000 UK adults, we report a number of novel findings: (1) the frequency of severe COVID-19 cases was higher in participants with CKD compared to those without CKD, regardless of ethnicity and socioeconomic status; (2) in those with CKD, accounting for potential confounders, Black and South Asian ethnic groups were around twice as likely to have severe COVID-19 compared to White participants and the most socially deprived groups were at 50%-60% increased risk of severe COVID-19; (3) associations of ethnicity or deprivation with severe COVID-19 were similar, regardless of the presence of CKD.

Whilst previous studies have shown that CKD is associated with an increased risk of hospitalisation and death from COVID-19 [7, 13, 14], to our knowledge no previous data has explored how this risk differs across ethnic and socioeconomic groups in relation to CKD. Our results are consistent with the stark ethnic inequalities evident in recent studies of

COVID-19 mortality and outcomes [3, 4], including an analysis of 17 million people in England using the OpenSAFELY platform [5]. We found that the likelihood of severe COVID 19 was increased in the most deprived CKD socioeconomic groups. This finding supports previous reports in the general population [6, 7, 9-11]. Notably, as we did not find evidence that associations for ethnicity and socioeconomic status were modified by presence of CKD, the relative risk of severe COVID-19 was similar in participants with and without CKD and the presence CKD similarly increased the relative risk irrespective of ethnicity or deprivation.

Many explanations exist as to why there may be an elevated risk of SARS-CoV-2 infection in minority ethnic or socially deprived groups. Possible reasons include living in deprived areas; working in high-exposure or front-line occupations; living in large, multigenerational households; a higher burden of comorbidity; discrimination; and poor access to health services [3, 5, 9].

There are limitations to this analysis. The cohort consists of volunteers, meaning results may not be entirely generalisable to the whole UK [18]. We used data, particularly CKD classification, from the baseline assessments recorded >10□years ago and, as such, the current health status of participants (e.g., changes in kidney function) cannot be determined. Our CKD group had mild disease staging and findings may differ as disease severity increases. We were only able to categorise ethnicity into three broad categories, rather than considering disaggregated groupings. Disaggregation into specific ethnicity groups has revealed differences in COVID-19 outcomes [5].

This study is the first to detail the association of ethnicity and socioeconomic deprivation with the risk of severe COVID-19 in relation to presence of CKD. These findings are of importance in informing interventions to reduce morbidity and mortality amongst these groups and policy and practice improvements are needed to address the broad disparity among CKD patients.

## Data Availability

Data available through UK Biobank.

## Acknowledgments

KK is supported by the National Institute for Health Research (NIHR) Applied Research Collaboration East Midlands (ARC EM) and the NIHR Leicester Biomedical Research Centre (BRC). TY is funded by a grant from the UKRI-DHSC COVID-19 Rapid Response Rolling Call (MR/V020536/1).

