## Supplementary material for "Association of chronic kidney disease, ethnicity and socioeconomic status with COVID-19 hospitalisation and mortality: a UK Biobank study"

**Online supplementary material**

**Supplementary material 1. Number of COVID-19 positive inpatient tests, deaths, and severe COVID-19 across ethnic and socioeconomic deprivation groups in subjects with and without CKD**

|  | **Positive inpatient tests** | | **Death from COVID-19** | | **Severe COVID-19** | |
| --- | --- | --- | --- | --- | --- | --- |
|  | **CKD** | **Non-CKD** | **CKD** | **Non-CKD** | **CKD** | **Non-CKD** |
| Total | 169 (1.6%) | 4338 (1.0%) | 78 (0.7%) | 953 (0.2%) | 247 (2.4%) | 5291 (1.2%) |
| *Ethnicity* | | | | | | |
| White | 146 (1.5%) | 3905 (0.9%) | 67 (0.7%) | 873 (0.2%) | 213 (2.2%) | 4778 (1.1%) |
| Black | 12 (3.5%) | 120 (1.8%) | 6 (1.7%) | 29 (0.4%) | 18 (5.2%) | 149 (2.2%) |
| South Asian | 7 (3.5%) | 181 (2.1%) | 3 (1.5%) | 32 (0.4%) | 10 (5.1%) | 213 (2.5%) |
| *Socioeconomic deprivation status* | | | | | | |
| Least deprived | 59 (1.1%) | 1840 (0.8%) | 30 (0.6%) | 366 (0.2%) | 89 (1.7%) | 2206 (0.9%) |
| Average | 67 (2.0%) | 1478 (1.0%) | 21 (0.6%) | 325 (0.2%) | 88 (2.7%) | 1803 (1.3%) |
| Most deprived | 43 (2.2%) | 1010 (1.4%) | 27 (1.4%) | 262 (0.4%) | 70 (3.5%) | 1272 (1.8%) |

Data shown as n (%) for all n=459,042 individuals classified as CKD or not (i.e., availability of creatinine value)

CKD = Chronic kidney disease

**Supplementary material 2. Association of ethnicity and social deprivation index with severe COVID-19, in subjects with and without CKD**

|  | **OR** | **Upper 95%CI** | **Lower 95%CI** | **P-value** |
| --- | --- | --- | --- | --- |
| **CKD (n=9935)** |  |  |  |  |
| *White (reference)* | 1.000 | - | - | - |
| Black | 2.256 | 1.338 | 3.803 | 0.002 |
| South Asian | 2.065 | 1.065 | 4.004 | 0.032 |
| *Least deprived (reference)* | 1.000 | - | - | - |
| Average | 1.497 | 1.099 | 2.039 | 0.011 |
| Most deprived | 1.625 | 1.145 | 2.306 | 0.007 |
| **Non-CKD (n=432,525)** |  |  |  |  |
| *White (reference)* | 1.000 | - | - | - |
| Black | 1.625 | 1.369 | 1.930 | <0.001 |
| South Asian | 1.908 | 1.653 | 2.203 | <0.001 |
| *Least deprived (reference)* | 1.000 | - | - | - |
| Average | 1.289 | 1.208 | 1.375 | <0.001 |
| Most deprived | 1.661 | 1.542 | 1.789 | <0.001 |

Data shown for all individuals classified as CKD or not with data for confounding variables

Data presented as odds ratio (OR) and 95% confidence intervals (95%CI)

Adjusted for current age, sex, obesity, number of cancer and non-cancer illnesses (cancer illnesses included any incidence of cancer including bowel, skin, prostate, and leukaemia; non-cancer illnesses included cardiovascular disease, respiratory conditions, diabetes, and neurodegenerative disease)

CKD = Chronic kidney disease
